# Material properties of urethral stents for hypospadias repair

**DOI:** 10.1101/2021.04.06.21254967

**Authors:** Tawan Jamdee, Christopher Foster, Courtney K. Rowe, Kelly A. Burke

**Author notes:** **Corresponding Author**: Kelly A. Burke, Ph.D., Assistant Professor, *University of Connecticut*, Chemical and Biomolecular Engineering and Polymer Program, Institute of Materials Science Room 205B, Storrs, CT 06269-3136.

## Abstract

**Introduction:** Despite the prevalence of hypospadias surgery and the near ubiquitous use of postoperative urethral stents, there has been no evaluation of the material properties of commonly used choices. Our study sets out to close this gap with an evaluation and comparison of the material properties of four urethral stents commonly used after hypospadias surgery.

**Study Design:** Thermal analysis and mechanical analysis of the Zaontz Urethral Stent, the Firlit-Kluge Urethral Stent, the Koyle Diaper Stent, and the Bard Premature Infant Feeding Tube were performed.

**Results:** Thermal analysis shows that all four compositions tested are rubbery polymers at body temperature, with glass transition temperatures far below human body temperatures. The Zaontz and Koyle stents are thermoplastic elastomers with strong melting transitions above body temperature, but the Firlit-Kluge stent is completely amorphous at body temperature and is likely chemically cross-linked to generate the polymer network. The Bard feeding tube was by far the stiffest, with a Young’s Modulus of 14.0± 0.78 (compared to the Zaontz stent at 4.12 ± 0.56, the Firlit-Kluge stent at 4.92± 0.63, and the Koyle stent at 4.09± 0.49.) The Firlit-Kluge stent was the strongest, with 84.3± 2.83 MPa required to fracture it compared to the Zaontz stent at 65.5 ± 2.57, the Koyle stent at 66.8± 3.16, and then Bard feeding tube at 34.6± 1.89.

**Discussion:** While there is little information associating urethral stent type with outcomes after hypospadias surgery, material properties may account for findings of prior studies. Stiffer stents may contribute to decreased postoperative comfort, while a stent that is too soft and extensible may have issues with dislodgement, kinking and breaking.

**Conclusion:** This study provides a foundation for future work optimizing urethral stents, designing support for regenerative medicine applications, and improving hypospadias outcomes.

## 1. Introduction

Hypospadias impacts an estimated 1 in 250-350 U.S. male births.^1^ Nearly all will undergo surgical repair, and nearly all repairs utilize a urethral stent postoperatively.^2^ Yet little is known about the material properties of commonly used urethral stents. This gap in the literature prevents future work to optimize stent composition and design. This may impact surgical and patient-centered outcomes as well as future regenerative medicine techniques, all of which rely on urethral stent use. Our study sets out to close this gap with an evaluation and comparison of the material properties of four urethral stents commonly used after hypospadias surgery.

## 2. Material and methods

### 2.1. Materials

Four stents commonly used after hypospadias surgery at our independent children’s hospital were purchased directly from the manufacturer (**Table 1**). Materials are as noted on the manufacturer website or patent. Estimated cost per stent is an average rounded to the nearest dollar based on two separate quotes from hospital systems with contracts with the manufacturer.

**Table 1.**
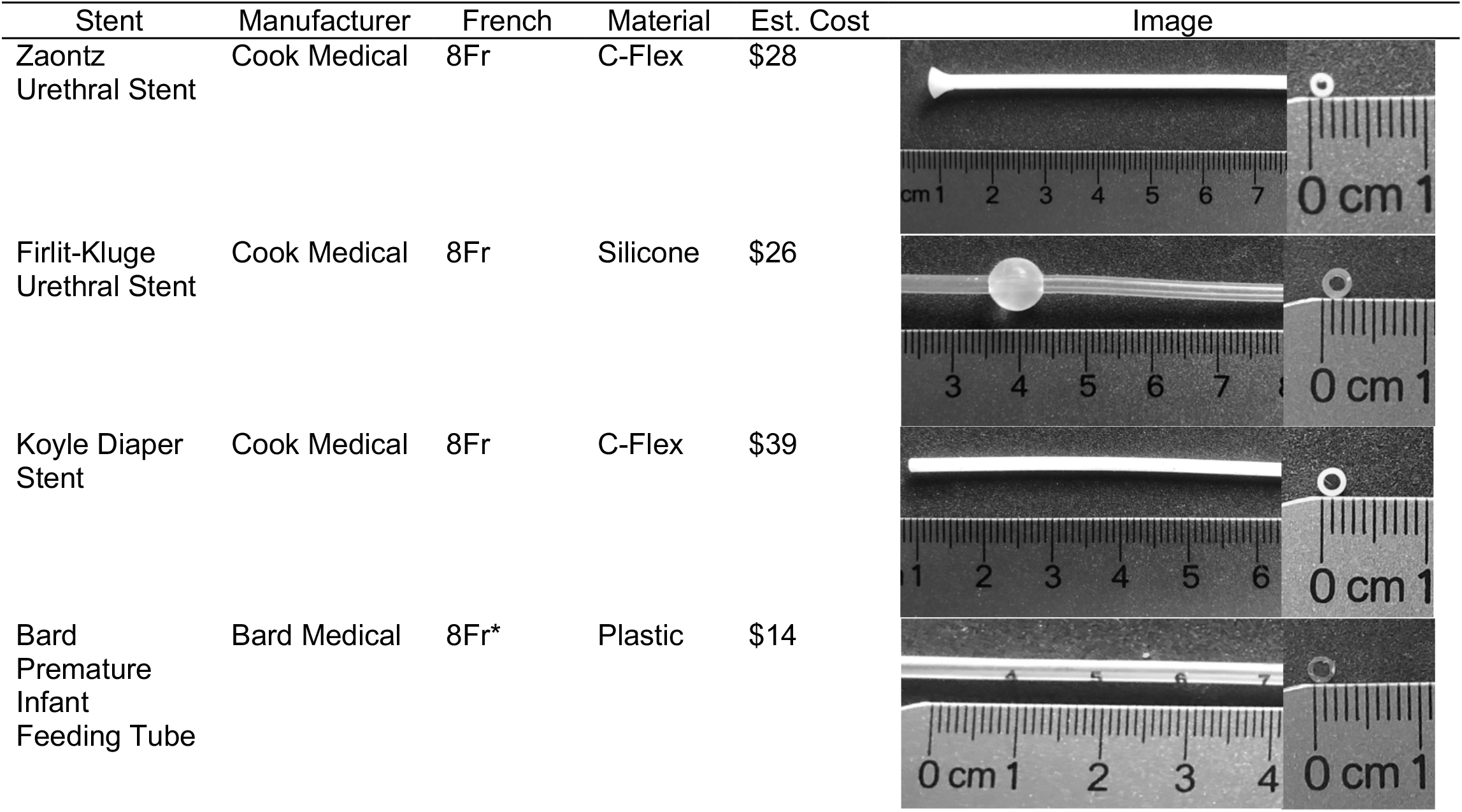
Overview of evaluated hypospadias stents.

| Stent | Manufacturer | French | Material | Est. Cost | Image |
| --- | --- | --- | --- | --- | --- |
| Zaontz Urethral Stent | Cook Medical | 8Fr | C-Flex | \$28 | |
| Firlit-Kluge Urethral Stent | Cook Medical | 8Fr | Silicone | \$26 | |
| Koyle Diaper Stent | Cook Medical | 8Fr | C-Flex | \$39 | |
| Bard Premature Infant Feeding Tube | Bard Medical | 8Fr* | Plastic | \$14 | |

### 2.2. Thermal Analysis

The thermal properties of the stent materials were investigated because the location of the polymer’s thermal transitions relative to room temperature and body temperature will impact the mechanical behavior of the polymers. Uniform samples were cut from each stent so that the samples lacked non-polymeric components, such as inks. Thermogravimetric Analysis (TGA) was run on 8-12 mg samples. Samples were placed in titanium pans and heated from 25 °C to 800 °C at a rate of 10 °C min^-1^ under flowing nitrogen atmosphere. The threshold for thermal degradation was set as the temperature where the sample lost 1% of its original weight. All samples were tested in a TA Instruments TGA Q500 (New Castle, DE) controlled by Q Series Software, and data were analyzed using TA Instruments Universal Analysis 2000.

The thermal transitions of a polymer are impacted by its composition as well as its processing history. Differential Scanning Calorimetry (DSC) can be used to characterize the polymer’s glass transition and melting and recrystallization transitions. The first heating trace was studied because it informs about the state of the polymer when it is in the stent and as it will be used in the patient. To compare materials from different stents, it is necessary to compare materials at the same thermal history, so for this reason the second heating and cooling cycles were also acquired. Uniform sections of stents were cut into 12-14 mg samples and encapsulated in aluminum pans. Samples were run in a TA Instrument DSC Q20 (New Castle, DE) using the following thermal program: heat to 200 °C at a rate of 10 °C min^-1^, hold isothermally for 5 min, cool to −70 °C at 10 °C min^-1^, and then hold isothermally for 5 min. The cycle was then repeated twice, with the thermal transitions measured on the first cycle to capture processing effects and the second cycle to compare materials with the same thermal history. Data were analyzed using TA Instruments Universal Analysis 2000. The temperature of the glass transition was measured from the midpoint of the stepwise change in heat flow on the heating trace, and the temperatures of the melting and crystallization transitions being taken from the heat flow trace as the minimum of an endothermic valley or maximum of an exothermic peak, respectively. Latent heats of melting and crystallization were quantified by measuring the area of the peak.

### 2.3. Mechanical Analysis

Dynamic Mechanical Analysis (DMA) was performed to determine the mechanical properties of the stents. Young’s Modulus is a property that quantifies the resistance of the polymer to deformation prior to the onset of plastic deformation. Storage modulus and loss modulus are dynamic mechanical properties used to characterize elastic and viscous character of the polymers, respectively. Polymers are viscoelastic materials, which means that they respond to applied stress (or strain) with properties that behave like both an elastic solid and a viscous liquid.

Young’s modulus, storage modulus, and loss modulus were measured using a TA Instruments Q800 Dynamic Mechanical Analyzer (DMA) (New Castle, DE, **Figure 1a**) controlled using Q Series Software. Rectangular samples (typical size: 20 mm length by 7 mm width by 250 μm thickness) were cut from sections of stents that were uniform and lacking in holes or curvature. To measure Young’s modulus, samples were loaded into the tension film clamp with a 0.1 N preload force, and force was ramped at a rate of 0.5 N min^-1^ up to a maximum of 18 N. The sample deforms in response to applied force, and length is measured by the instrument. Strain (ε) is expressed in percent and is calculated by dividing the sample’s measured length (*L*) by its length when it was loaded with the preload force (*L*_*o*_) using Eqn. 1:

**Figure 1.**
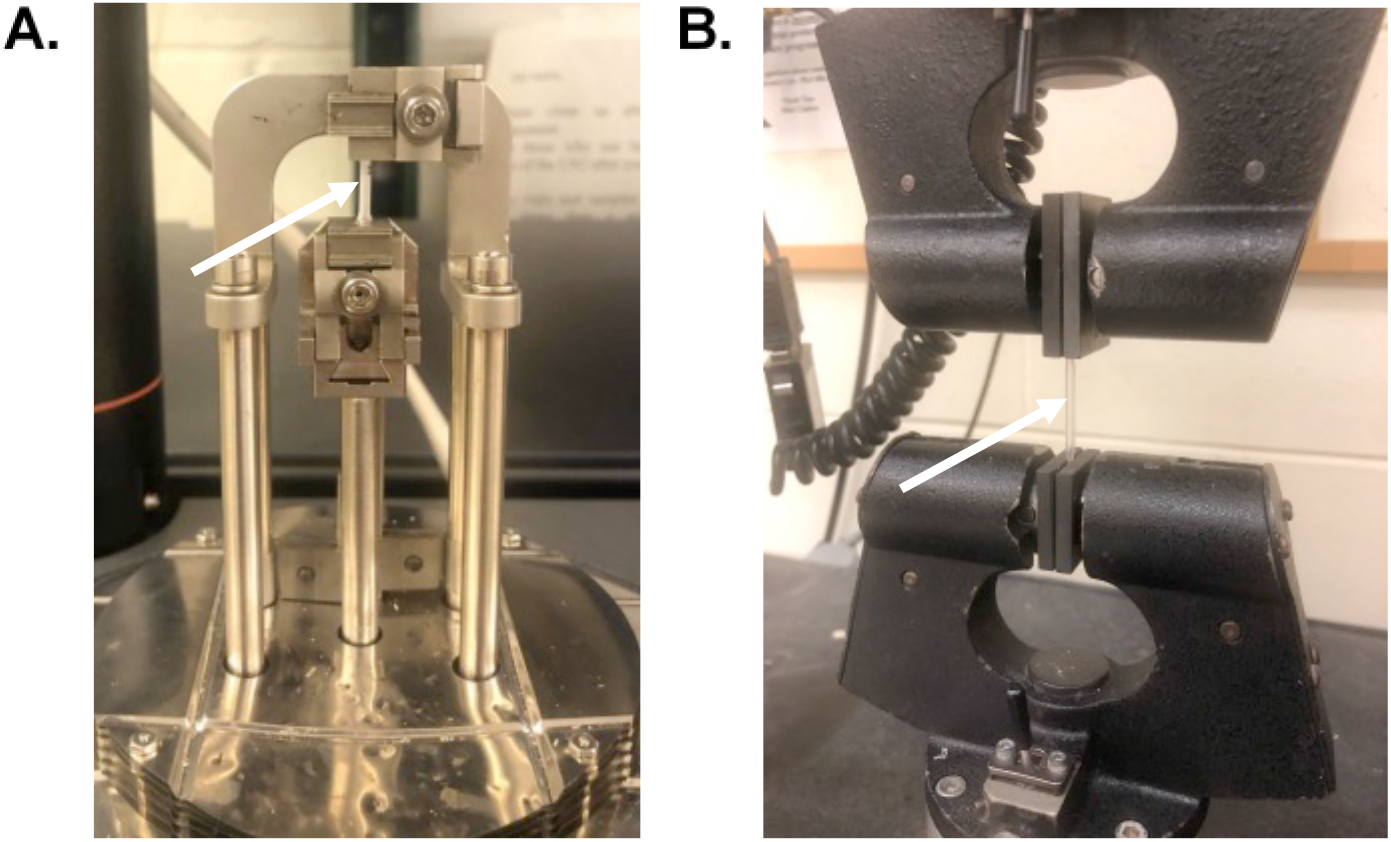
Stent sample loaded in tension within the clamps of the (A) Dynamic Mechanical Analyzer and (B) Instron. For both images, the placement of the tensile specimen is noted by the white arrow.

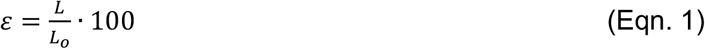

Young’s modulus was determined from the initial slope of the stress versus strain curve, where strain was less than 3%. A minimum of five replicates were tested for each stent. To measure storage and loss moduli, films were cut from stents as described above and loaded into the tension film clamp with a preload force of 0.1 N. The materials were oscillated with a strain of 2% at a frequency of 1 Hz at 25 °C. As urethral stents may be subjected to repeat episodes of extension and contraction, a cyclic loading-unloading study was performed using the DMA. The cyclic tests were conducted in tension by mounting films cut from the stent in tension in the DMA, loading the sample by increasing strain of the sample at a rate of 3% min^-1^ until reaching to 100% strain, unloading the stent by decreasing strain at the same rate until reaching 1% strain, and repeating the cycle two additional times.

To gain a more complete description of stent extensibility, ultimate stress and strain were characterized to determine the stress and strain, respectively, that stent could withstand before fracture. Ultimate properties were measured on an Instron 5869 (Norwood, MA, **Figure 1b**) controlled by Bluehill 2 (Glenville, IL). Uniform rectangular film samples were cut from stents as described above (typical size: 20mm length by 7mm width by 250 μm thickness). Samples were loaded into tension clamps before stretching at a rate of 50 mm min^-1^ until failure at 25 °C. Ultimate Stress was determined to be the stress at a fracture and ultimate strain was determined to be the strain at fracture.

## 3. Results

Thermogravimetric analysis (TGA) was used to characterize stent degradation and shows that all four stent compositions tested degrade well above body conditions (**Figure 2a,c**). All four compositions of stents measured by DSC (**Figure 2b,d**) have subambient glass transition temperatures, meaning that all stents are in a rubbery (nonglassy) phase at room temperature and body temperature. The Zaontz Stent and the Koyle stent display very similar melting temperatures and latent heats of fusion upon heating, indicating similar types and amounts of crystallinity, though wide-angle x-ray analysis was not used to characterize crystal forms (**Figure 2d**). Similarly, cooling the Zaontz stent and the Koyle stent results in similar observed temperature and latent heats of crystallization. Because both Zaontz and Koyle stents display crystalline transitions above 37 °C, these polymers function as thermoplastic elastomers when implanted in the body, where the polymer chains are soft and rubbery but the crystals prevent the materials from flowing when stress is applied.

**Figure 2.**
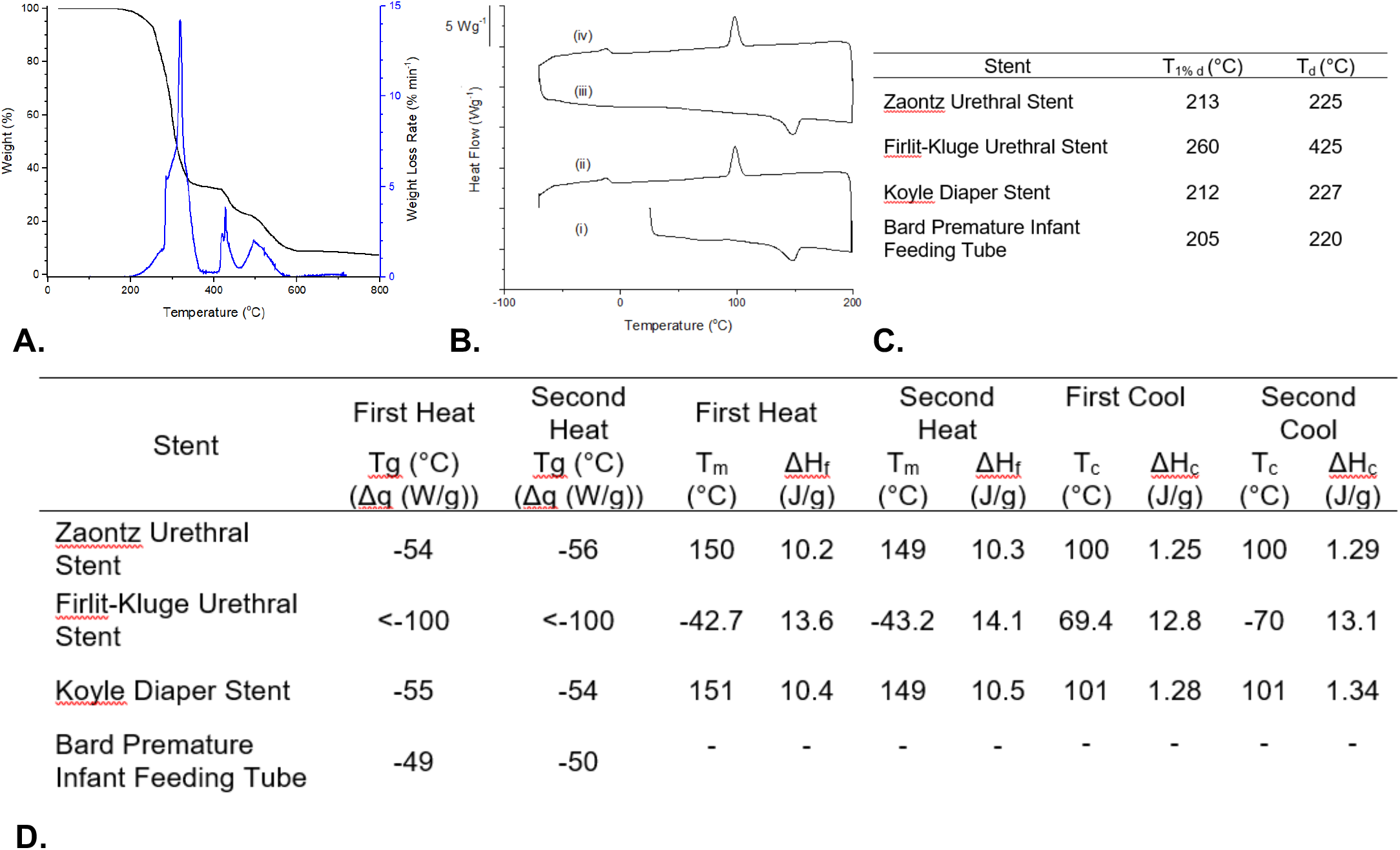
Thermal properties of urethral stent materials. (A) Thermogravimetric analysis of Zaontz Urethral Stent upon heating at 10 °C min^-1^ under flowing nitrogen atmosphere, where the black trace denotes the weight percent relative to original sample weight (units: %) and blue trace is the derivative of the weight loss with respect to time (units: % min^-1^). (B) First heating (i), first cooling (ii), second heating (iii), and second cooling (iv) of Zaontz Urethral Stent at a rate of 10 °C min^-1^ using differential scanning calorimetry (DSC). (C) Thermal decomposition of stenting materials as measured using thermogravimetric analysis, where T_1%d_ is the temperature where 1% weight loss has occurred and T_d_ is the temperature at which the rate of weight loss first exceeds 1% min^-1^. (D) Stent thermal transitions stents measured using DSC, where Tg denotes the glass transition, T_m_ is the melting temperature, and T_c_ is the crystallization temperature. Latent heats of fusion (ΔH_f_) and crystallization (ΔH_c_) transitions are quantified by calculating the area of the peak associated with the transition.

In contrast, the Firlit-Kluge stent does not display a glass transition temperature over the testing range (**Figure 2d**), but this is to be expected because of the low glass transitions associated with silicone polymers. The Firlit-Kluge stent displays a melting peak upon heating around −43 °C and the polymer crystallizes around − 70 °C upon cooling, but no other transitions were observed. These results indicate the Firlit-Kluge is a rubbery elastomer at room temperature and body temperature. Additionally, the lack of additional transitions suggest that the Firlit-Kluge is chemically cross-linked, which would permit the stent to resist flowing without crystalline content at body temperature. Chemical cross-links serve a similar mechanical function as the crystallites of thermoplastic polymers, however materials that are chemically cross-linked are not reprocessable. Attempts to dissolve the Firlit-Kluge stent in common solvents, including tetrahydrofuran, chloroform, and toluene proved unsuccessful to further support that this material is chemically cross-linked.

The Bard Premature Infant Feeding Tube has a subambient glass transition and displays rubbery behavior at 37 °C. The Bard tube also displays a small endotherm on the first heat around 70 °C, but this is not present on subsequent heating traces and no crystallization is observed. It is possible that the transition at 70 °C is a melting transition, that permits the material to resist flowing under applied stress at 37 °C. This transition may also be due to plasticizer within the material, but additional modulated DSC failed to provide a definitive answer. The Bard stent is not chemically cross-linked, as it does flow when temperature is raised and it will dissolve in tetrahydrofuran, therefore the transition at 70 °C is expected to be critical for how this stent functions as a thermoplastic elastomer.

**Figure 3a** shows a representative stress strain curve of a Bard feeding tube, where the Young’s modulus was found by measuring the slope within 3% of the initial strain. The Young’s modulus for Bard Premature Infant Feeding Tube is about 14 MPa, but the elastic modulus is about one third of this value for the other three compositions. (**Figure 3b**) Storage modulus exceeds loss modulus for all compositions, demonstrating that all materials display more solid-like behavior than liquid-like behavior (**Figure 3c**). There are no significant differences between the Zaontz, Firlit-Kluge, or the Koyle stents, but the Bard stent again displays a significantly higher storage modulus than the other materials. These results indicate that the Bard stent displays a significantly higher stiffness than the other stents, which may impact patient comfort. Finally, the Zaontz, Firlit-Kluge, or the Koyle stents all display similar loss tangent values, which is the ratio of loss modulus to storage modulus and is indicative of how well a material dissipates energy. The loss tangent is highest for the Bard stent at 0.35, which is about double the values measured for the other stents and may be due to the proximity of the testing temperature (25 °C) to the material transition that occurs around 70 °C.

**Figure 3.**
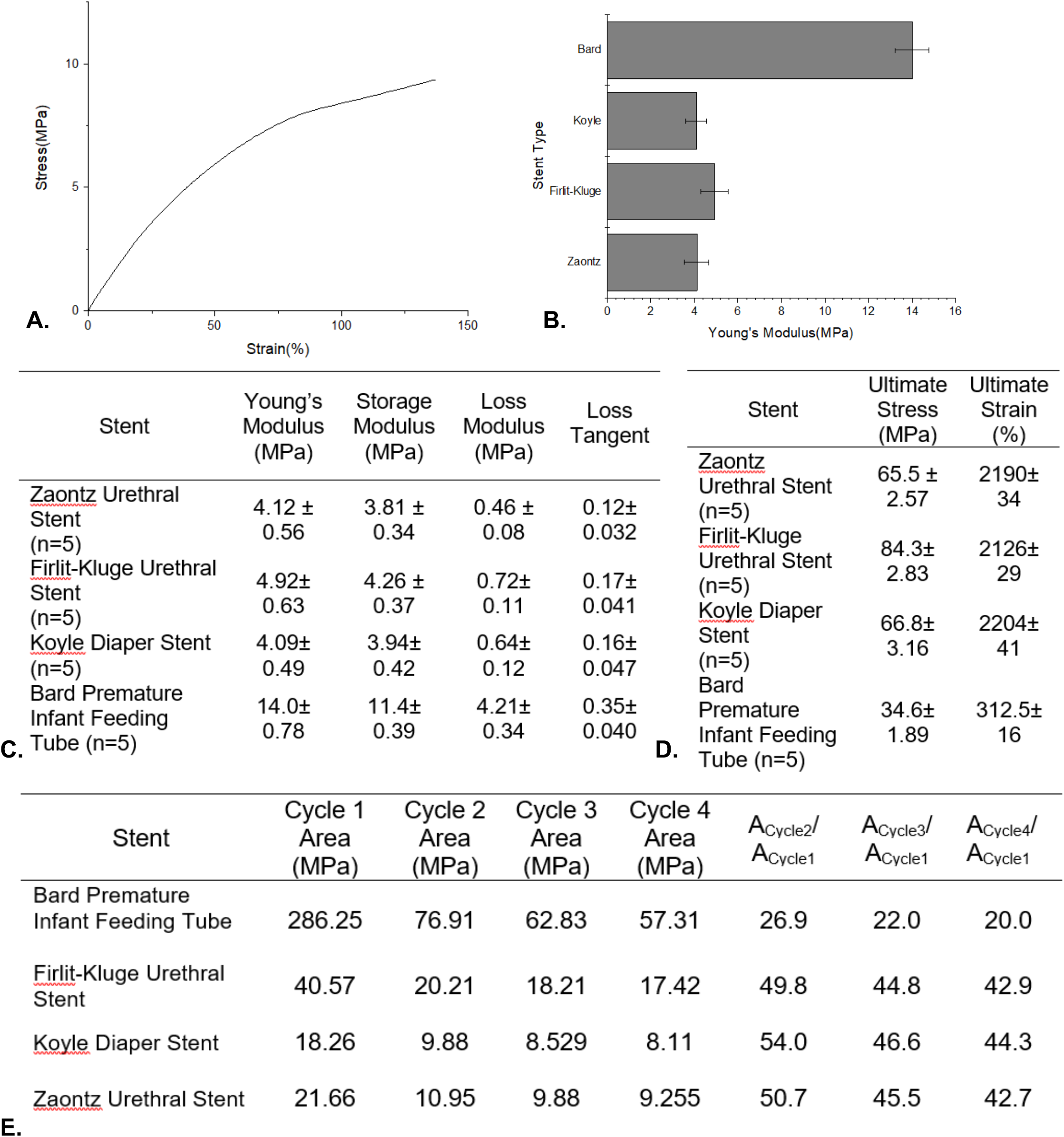
Mechanical properties of urethral stenting materials. (A) Example stress-strain curve of Bard feeding tube used to determine mechanical properties of the stents. Stents were stretched by ramping tensile force from 0.1 N to 18 N at 25 °C. (B) Young’s Modulus of stents calculated from the stress-strain curves measured by the DMA. (C) Summary of dynamic mechanical properties collected on stents using DMA at 25 °C. (D) Summary of Ultimate properties measured using the Instron at displacement rate of 50 mm min^-1^ and at 25 °C, where ultimate stress and ultimate strain were obtained from the point of fracture. (E) Summary of cyclic loading and unloading properties measured on stents using the DMA at 25 °C.

The Zaontz, Firlit-Kluge, and the Koyle stents are all highly extensible, deforming to more than 2000% strain before breaking. The Zaontz and the Koyle stents fracture at a similar stress, but the Firlit-Kluge was observed to require a significantly higher amount of applied stress to fracture, which may be due to its cross-linked network structure that gives rise to a stronger material. The Bard stent was found to be the highest stiffness, the least extensible, and to fracture at the lowest applied stress (**Figure 3d**).

All stents show the ability to be deformed through cyclic loading and unloading cycles (**Figure 3e, Supplemental Figure 1**.). The area reported in **Figure 3e** is calculated by subtracting the area under the loading and unloading curves as shown in Eqn. 2, and this value represents the energy that is lost upon cycling the material.

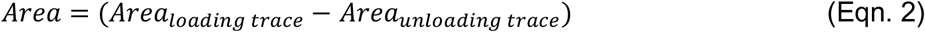

The ratio of area relative to the area from the first cycle is also provided in **Figure 3e**. All stenting materials show the typical trends on loading and unloading studies, where the first cycle has the largest unrecovered area and all following cycles display much smaller changes. The Bard Feeding Tube displayed the largest first load area amongst all other stents, however also showed the greatest loss in area with subsequent loading. The Zaontz and Koyle stent show similar values in **Figure 3e** and the shape of their curves shown in **Supplemental Figure 1** also further support them being the same material.

## 4. Discussion

This paper is the first to describe and compare material properties of commonly used hypospadias stents. Our study found that all stents have glass transition temperatures below human body temperature. The thermal properties of the Zaontz and Koyle stents were nearly identical, as expected since both are made out of the proprietary material C-Flex. The Firlit-Kluge stent’s thermal properties indicate that this material is likely a chemically cross-linked silicone, rather than a thermoplastic like the other stents. The Bard feeding tube results were slightly surprising because this material has a lower amount of crystallinity than the other stents, yet the mechanical properties of the Bard stent show a significantly higher stiffness and lower extensibility. One possible explanation is that the Bard stent may be formed from a polymer with a more rigid backbone or whose side chains hinder polymer chain rotations, which can account for the higher elastic modulus even though this material does not crystallize as readily as the other materials. There were small differences in stiffness and extensibility of the Zaontz, Koyle, and the Firlit-Kluge when it comes to material stiffness and extensibility, however the Firlit-Kluge appears stronger because it fractures at a higher stress.

Urethral stent use after hypospadias surgery remains a controversial topic. A number of metanalyses and prospective studies have been unable to demonstrate a significant difference in fistula or stenosis rates after either distal repair or hypospadias fistula repair.^3-5^ Despite this, the majority of surgeons continue to use urethral stents,^2^ perhaps because of their benefit in preventing the estimated 1-12% rate of urinary retention after surgery.^6,7^ To date, there is no high quality evidence associating type of urethral with any significant difference in long term hypospadias surgery outcomes.

Some studies have suggested that certain stents are more comfortable for patients. Ozcan et al performed an unblinded, randomized study to compare the Zaontz stent to an infant feeding tube like the Bard tube evaluated in our study.^8^ Patient comfort was assessed using the Face, Legs, Activity, Cry, Consolability (FLACC) scale and was higher for the Zaontz stent on the third postoperative day only. This difference in comfort may have been due to the difference in stiffness of these two stents, as demonstrated in our study by their different Young’s modulus (4.12 for the Zaontz stent vs. 14.0 for the Bard Feeding Tube), but stent design and placement may have also played a role. The Zaontz stent has a flange that sticks only a small way out of the meatus, while the feeding tube is longer and therefore can move more, which may increase discomfort. The Zaontz stent was also placed just within the external sphincter, while the feeding tube was placed within the bladder. The spasms associated with stents placed within the bladder have been noted on a number of studies, and many have contributed to the results in the Ozcan study.^6^

Lee et al. compared the Koyle stent to a stent made of modified silastic tubing in a prospective, non-randomized fashion.^9^ Silastic is a proprietary name for a type of flexible, transparent silicone, which we can assume is similar in material properties to the silicone Firlit-Kluge stent tested in our study. There was an increase in unplanned visits to the emergency room for the silastic tubing group, primarily because the stents fell out early. In part this was due to a difference in placement technique. The silastic stents were secured with a nonabsorbable suture and planned to be removed in clinic, so if they became dislodged the parents were distressed. The Koyle stents were placed with an absorbable suture and expected to fall out on their own, so no emergency room visit was needed if this happened early. Unfortunately, the study authors did not record how long each stent remained in place and whether early dislodgement was higher for the silastic stent group then the Koyle stent group. However, there is a possible material properties explanation for issues of dislodgment, tearing and kinking noted by Lee et al with the silastic stent. The Firlit silicone stent evaluated in our study was soft and extensible. It is possible that while a soft stent may be better for patient comfort, a stent that is too soft and extensible is at risk of kinking, tearing, and dislodgement in clinical practice if these materials limitations are not accounted for with modifications in stent design.

It is interesting to compare results from this study to the work performed by Cunnane et al, who recently evaluated the mechanical properties of donor urethral tissue as a foundation for future tissue engineering work.^10^ The donor material tested in that study resulted in much lower modulus than the urethral stents tested here, making it less stiff than any commonly used stent. This was expected, but it is useful to note that care should be taken when comparing the two studies because the testing methods do differ. Additionally, the impact of isolated urethral testing outside the mechanical support of the penis is unclear, as is the impact of estrogen on the tissue as the samples were taken during male to female gender reassignment surgery and patients can be presumed to have been received gender affirming hormones prior to donation.

The main limitation of this paper is that the materials properties were evaluating while controlling for size and shape of the stent. This is why, despite the relative similarities in material properties of the Zaontz, Firlit-Kluge, and Koyle stents, each feels different when being handled. How a stent can be used by a surgeon and how it feels to a patient will depend also on its shape and design, which were not evaluated here.

## 5. Conclusions

This study shows that there is a wide margin of acceptable stiffness and extensibility of stents without only minimal evidence of impact on surgery outcomes or patient quality of life. The only similarity across all stents is the thermal stability at body temperature.

In the absence of clear clinical differences, it is interesting to consider the cost variation between the stents, as the Bard feeding tube is significantly cheaper (**Tab. 1**) The final selection of a stent will ultimately be based on surgeon preference. This study provides data that can be used for future improvements in stent design.

## Data Availability

If requested, data and material samples will be made available for sharing to qualified parties as determined by the PI, as long as such a request does not compromise intellectual property interests, interfere with publication, or precede data curation.

## SUPPLEMENTAL TABLES AND FIGURES

**Supplemental Figure 1.**
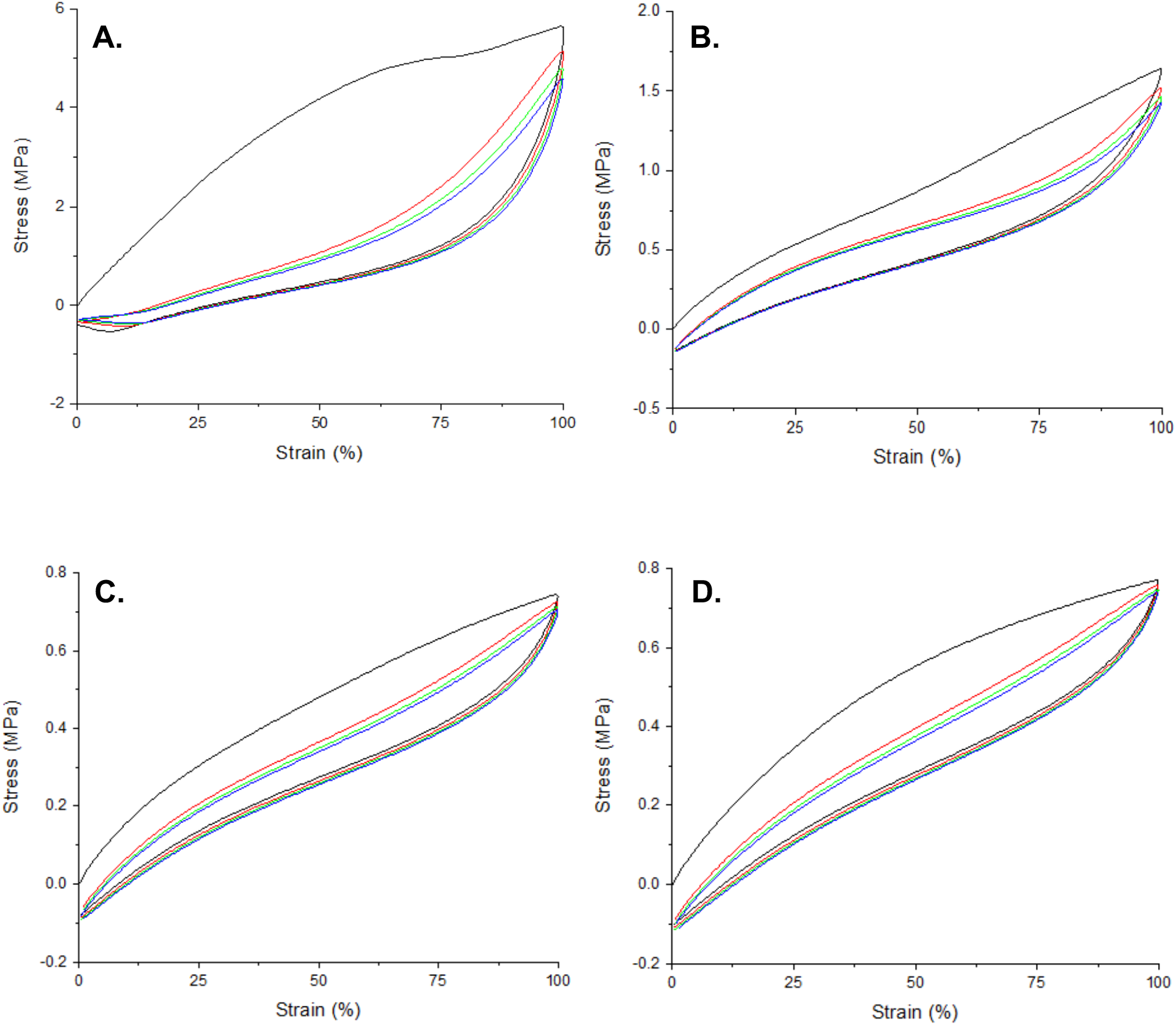
Cyclic loading and unloading plots of (A) Bard Feeding Tube, (B) Firlit-Kruger Stent, (C) Koyle Stent, and (D) Zaontz. All samples stretched to 100% deformation at 25 °C for 4 cycles. Black trace denotes first cycle, red trace denotes second cycle, green trace denotes third cycle, and blue trace denotes fourth cycle.

